# NIRDuino: A modular, Bluetooth-enabled, Android^®^-configurable fNIRS system with dual-intensity mode built on Arduino^®^

**DOI:** 10.1101/2024.12.20.24318425

**Authors:** Anupam Kumar, Seth Crawford, Tiffany-Chau Le, Ali Rahimpour Jounghani, Laura Moreno Carbonell, Alexandra Sargent Capps, Alec B. Walter, Daniel Liu, Reed Sullivan, E. Duco Jansen, SM Hadi Hosseini, Audrey K. Bowden

## Abstract

**Significance:** We present NIRDuino: an Open-source Android^®^-configurable, modular, and Bluetooth-enabled fNIRS system that allows researchers to perform neuroimaging studies with up to eight emitters and 16 detectors. The complete system (including Android tablet) can be assembled for less than $1000, and the emitters and detectors can be arranged in any configuration to achieve the desired short and long channels required for their study.

**Aim:** The system has been designed with non-engineers in mind, and the researcher only needs to design the wearable interfaces to attach the emitters and detectors to the body appropriate for their intended application.

**Approach:** The system consists of a battery-powered, wireless controller built on the Arduino® Nano ESP32 platform, a dongle with sockets for each of the eight emitters and detectors that can be connected, and individual wired probes for emitters and detectors. In accompaniment, Arduino®-based firmware and an Android^®^ application have also been developed and provided. The selected emitters and detectors can be arranged in any desired configuration, and the emitters can be configured to output light with both regular intensities and low intensities to collect data for “long channels” with sufficient signal quality and “short channels” without saturation. This paper details the system’s design and characterization on phantom and two physiological experiences on a human.

**Results:** The easy-to-configure hardware/software system demonstrated stability in fNIRS measurements using a single emitter-detector pair placed on a phantom, and reproduced previously published outcomes for arterial cuff measurements on the forearm and a arithmetic experiment on the forehead.

**Conclusion:** The NIRDuino circuitry and software demonstrated modularity and usability for NIRS experiments, and this low-cost platform will provide researchers globally with an affordable fNIRS system to easily adopt and adapt for their unique experimental needs.

## 1. Introduction

Functional neuroimaging is a promising strategy to uncover important clinical insights into many psychological diseases and disorders (for ex. ADHD^1–7^, Alzheimer’s disease^8–10^, dementia^11,12^). An ongoing challenge for neuroimaging studies, however, is to identify brain-wide associations with sufficient statistical significance. Addressing this challenge requires engaging *thousands* of subjects ^13^ and collecting several hours of data per subject to glean reproducible individual-level inferences^14^. Unfortunately, existing neuroimaging tools are not poised to meet this challenge due to their high costs and usability constraints^15^. To expand the scale of data collection, the field needs more devices to routinely engage with more subjects.

Continuous-wave functional near-infrared spectroscopy (CW-fNIRS) is an established technology for functional neuroimaging whose portability endows it with great potential to collect ecologically valid neuroimaging data^16,17^. CW-fNIRS measurements have been found to be correlated with functional magnetic resonance imaging (the gold standard of functional neuroimaging)^18,19^. Numerous mobile-interfaced CW-fNIRS systems^20–22^ have been published recently for fNIRS data collection over the prefrontal cortex, the auditory cortex, or the occipital cortex. Unfortunately, many of these systems lack the accessibility and modular configurability needed to be usable across diverse experimental requirements, as different experiments may require unique fields of view over the brain and unique choices of the number and placement of emitters and detectors. Placement of emitters and detectors are often defined based on the cognitive functions that researchers wish to evaluate and is facilitated by various tools such as the fNIRS Optode Location Decider (fOLD)^23^ or ninjaCap^24^. An additional consideration is whether to use short (<20 mm) or long (>20 mm) emitter-detector separation channels: short channels are used to collect non-neurological signals such as scalp hemodynamics and motion artifacts, which are later regressed out from long channel data for removing noise and non-neurological signal^25–30;^ in contrast, long channels collect both neurological signals and the signals collected by the short channels.

The wireless nature of some CW-fNIRS systems makes them more amenable to diverse environmental contexts^31,32^; however, several commercially available wireless and modular CW-fNIRS systems (i.e., certain models offered by vendors such as NIRx and Artinis) can only interface with personal computers (PCs) such as laptops and desktops. This prevents them from harnessing the cost and portability advantages that mobile phones and tablets provide and restricts the scale at which data can be collected. Albeit recent PCs such as the Microsoft Surface have taken a more tablet-like form factor, these systems come at significantly higher price points than tablet computers which again constrain adoption ^33^. Commercial wireless products such as Obelab’s NIRSIT^15^, NewmanBrain’s Theia^33^, and Mendi^10^ provide mobile apps for data collection, but their lack of modularity restricts the types of experiments for which they can be used. Several researchers have published research-grade modular designs for fNIRS studies recently, but they have not been open-sourced for the community to adopt or adapt^34–37^, limiting accessibility.

In contrast, open-source technology proffers significant cost advantages, increased participation from researchers, standardization of data collection, and better evaluation of reproducibility of results^38^. Moreover, open-source technologies can enable other researchers and developers to repurpose or augment efforts already made previously (ex. electronics design, software design) towards new wearables with new form factors for neuroimaging. Among the limited open-source options, non-modular systems such as FlexNIRS^39^ and DIY-NIRS^40^ have opened the door to increased accessibility, but their lack of modularity prevents them from being used during experiments requiring different emitter-detector arrangements. NinjaNIRS^41^ is a modular open-source fNIRS system, but it does not have wireless communication and mobile interfacing yet.

Additionally, while researchers looking to design systems for new applications could benefit from existing open-sourced systems, the level of sophistication for use of these systems requires that interest researchers have the requisite engineering training to understand and adapt systems according to their unique requirements. The inaccessibility of the technology due to cost and skill- level requirements not only bottlenecks the total volume of data that may be collected but also risks underrepresentation and exclusion of certain global communities as both participants and researchers.

A summary of the systems discussed is shown in Table 1. Thus far, no published or commercial CW-fNIRS system has addressed the combined needs for affordability, mobile interfacing, and modularity that can satisfy diverse experimental requirements without major electronics redesign. To this end, this paper presents NIRDuino: a modular, wireless, Bluetooth-based CW-fNIRS system built on the Arduino® platform and configurable using an Android^®^ mobile app. The system comprises a controller that can perform measurements with up to 8 emitters and 16 light detectors, enabling configurations with as many as 128 channels. The controller can wirelessly connect to an Android^®^-based mobile device using a custom app from which the researcher can perform multiple functions: select/unselect the emitters/detectors being used in an experiment, adjust the output light intensity for each emitter, visualize the data received using a real-time plot, and store the data locally on the mobile device in a .csv file. Moreover, each emitter-detector pair measurement is repeated at two separate user-defined light intensities so that both short (<20 mm) and long (>20 mm) emitter- detector separations can be accommodated. The complete system can be self-assembled for less than $1000 per unit, compared with the tens of thousands of dollars most commercial systems cost (including mobile tablet). Hence, for a fraction of the price of currently marketed systems, researchers can adopt multiple NIRDuino systems to engage a much larger number of research subjects than previously feasible. All hardware and software design files and output binaries have been shared using an Open-Source Foundation (OSF) repository along with instructions to easily set up and use the equipment.

**Table 1:** Comparison of available systems

|  | Portable | Modular | Open Source | Wireless Operation | Mobile Operation |
| --- | --- | --- | --- | --- | --- |
| **NewmanBrain Theia**<sup>42</sup> | x |  |  | x |  |
| **Obelab NIRSIT**<sup>43</sup> | x |  |  | x | x |
| **Obelab NIRSIT Lite**<sup>44</sup> | x |  |  | x | x |
| **Mendi**<sup>45</sup> | x |  |  | x | x |
| **Artinis Oxymon**<sup>46</sup> |  | x |  |  |  |
| **Artinis Brite MkII**<sup>46</sup> | x | x |  | x |  |
| **Artinis PortaLite MkII**<sup>46</sup> | x |  |  | x | x |
| **NIRx NIRScout**<sup>47</sup> |  | x |  |  |  |
| **NIRx NIRSport2**<sup>47</sup> | x | x |  | x |  |
| **BIOPAC fNIRx03S Series**<sup>48</sup> |  | x |  |  |  |
| **BIOPAC fNIRx04C**<sup>48</sup> |  | x |  |  |  |
| **BIOPAC fNIRSx03M**<sup>48</sup> | x |  |  | x |  |
| **Cortivision Photon Cap**<sup>49</sup> |  | x |  | x |  |
| **g.Tec g.Nautilus**<sup>50,51</sup> | x |  |  |  |  |
| **NIRSOptix**<sup>51</sup> |  |  |  |  |  |
| **GowerLabs Lumo**<sup>52</sup> | x |  |  |  |  |
| **Seenel Medelopt Mobility**<sup>53</sup> | x |  |  | x |  |
| **Seenel Medelopt Infinity**<sup>53</sup> | x |  |  | x |  |
| **Shimadzu LABNIRS**<sup>54</sup> |  | x |  |  |  |
| **Hitachi ETG-4000**<sup>55</sup> |  | x |  |  |  |
| **ISS Imagent**<sup>56</sup> |  | x |  |  |  |
| **HEGduino**<sup>57</sup> | x |  | x | x | x |
| **OpenNIRS**<sup>58</sup> | x | x | x | x |  |
| **NinjaNIRS**<sup>59</sup> |  | x | x |  |  |
| **FlexNIRS**<sup>39</sup> | x |  | x | x |  |
| **DIY-fNIRS**<sup>40</sup> | x |  | x | x | x |
| **NIRduino** | x | x | x | x | x |

The combined mobile interfacing, modularity, and affordability of NIRDuino will empower many fNIRS and NIRS-based researchers to conduct experiments on more individuals for longer durations, which can help meet the clinical need to identify statistically significant brain-wide associations. Moreover, the ease of use and low barrier to entry for researchers with all backgrounds will allow many to design novel wearable neuroimaging systems using the provided electronics and software designs. We are confident this will open the door for new neuroimaging studies that could not be conducted with existing hardware tools, and result in larger volumes of data crucial for drawing clinically meaningful and actionable insights

## 2. Methods

### 2.1. System Overview and Workflow

A high-level concept diagram of the system design and workflow for NIRDuino is shown in Figure 1, which highlights its two aspects: hardware and software. The hardware consists of four types of subsystems: light delivery probes (aka emitters, up to 8), light detector probes (aka detectors, up to 16), a central controller (black square labeled “Ctr”), and a dongle circuit (black rectangle labeled “Dg”) with sockets to interface the controller with the emitter and detectors. The system is versatile yet simple: the researcher just needs to connect the emitters and detectors they wish to use (Figure 1a), design and affix an enclosure to arrange the emitters and detectors on the anatomical location of interest (Figure 1b), select/deselect the emitters and detectors they wish to utilize using checkboxes on the configuration app (Figure 1c), and collect, observe, and store the data from the desired emitter-detector pairs (Figure 1d). The nature of the custom enclosure should consider the target site of research interest. The controller, dongle, and probes can be held onto the body using a variety of approaches, including rigid 3D printed parts and soft, flexible silicone interfaces. Example enclosures mounting all elements on the head and forearm are presented later in this paper, but other approaches are also achievable.

**Figure 1:**
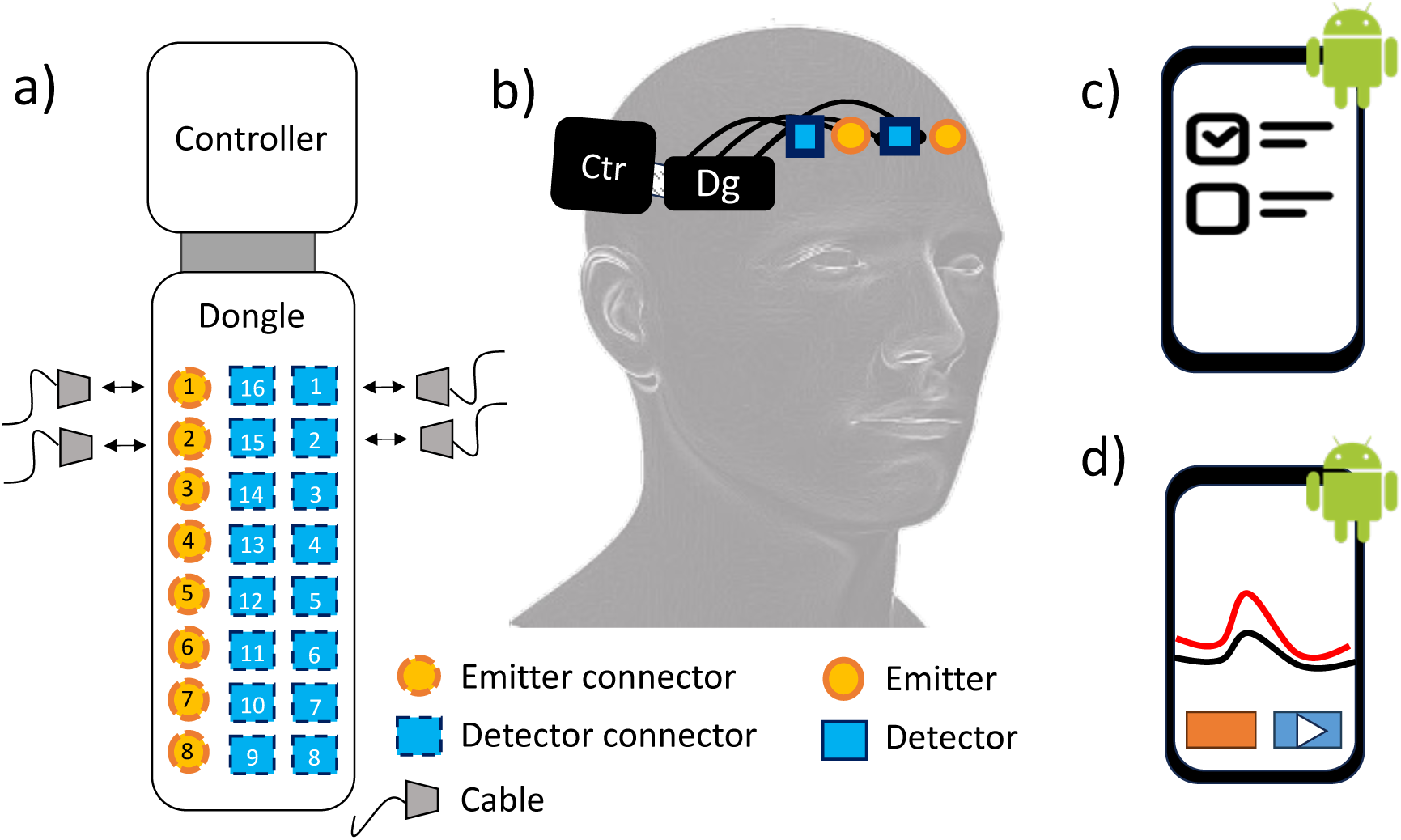
Concept diagram of the NIRDuino system and workflow. a) Step 1: Connect the desired emitters and detectors to the dongle. b) Step 2: arrange and anchor the emitters and detectors to the desired body part. The emitters and detectors connect to the dongle (“Dg”) which connects to the controller board (“Ctr”) using a flat flexible cable (FFC). c) Step 3: Select which emitters and detectors will be in use on the configuring mobile app. d) Step 4: collect, observe, and store the data on the Android® app.

### 2.2. Hardware Design

#### 2.2.1. Emitter design

Each emitter was designed to measure changes in oxy- and deoxy- hemoglobin and therefore consists of two light emitting diodes (LEDs) with wavelengths of 740nm (EPIGAP OSA Photonics) and 860nm (ams OSRAM USA Inc.). The chosen wavelengths are at either side of the isosbestic point of hemoglobin analytes at 808nm and minimize crosstalk during fNIRS measurements. Additional LED wavelengths for measuring Cytochrome-C Oxidase were also considered due to the recent rise in academic interest but were forgone due to insufficient proof of their value for neuroimaging purposes^60^. However, if new evidence emerges, these wavelengths may be included in future versions of this system.

Each LED is current-controlled using a 33-Ω series resistor and can be powered by an external variable voltage supply (VVS) with outputs ranging from 1.4V (minimum forward voltage) up to 5V, outputting light within the safety limitations set by the ANSI Z136.1-2014 and IEC 825-1 standards. Each LED is turned on or off using an NPN bipolar junction transistor (BJT) that can be toggled on/off using a digital signal. A schematic diagram describing this circuit is shown in Figure 2a. The controller (discussed later) supplies the VVS and toggle signals. The LEDs of each emitter are mounted onto the probes while the remaining part of the circuit (i.e., current-control resistors and NPN BJTs) is incorporated in the controller circuit (discussed later). Each emitter is miniaturized using 2-sided, 2-layer rigid printed circuit board (PCB) and has a 4-pin Japan Standard Terminal (JST) connector to interface with the controller. To minimize the size of the emitter, the BJT portion of the circuitry is integrated in the controller. The lateral dimensions of the emitter are 5.3 mm x 7.6 mm and its thickness is 7.7 mm. A photograph of the assembled emitter appears in Figure 2b.

**Figure 2:**
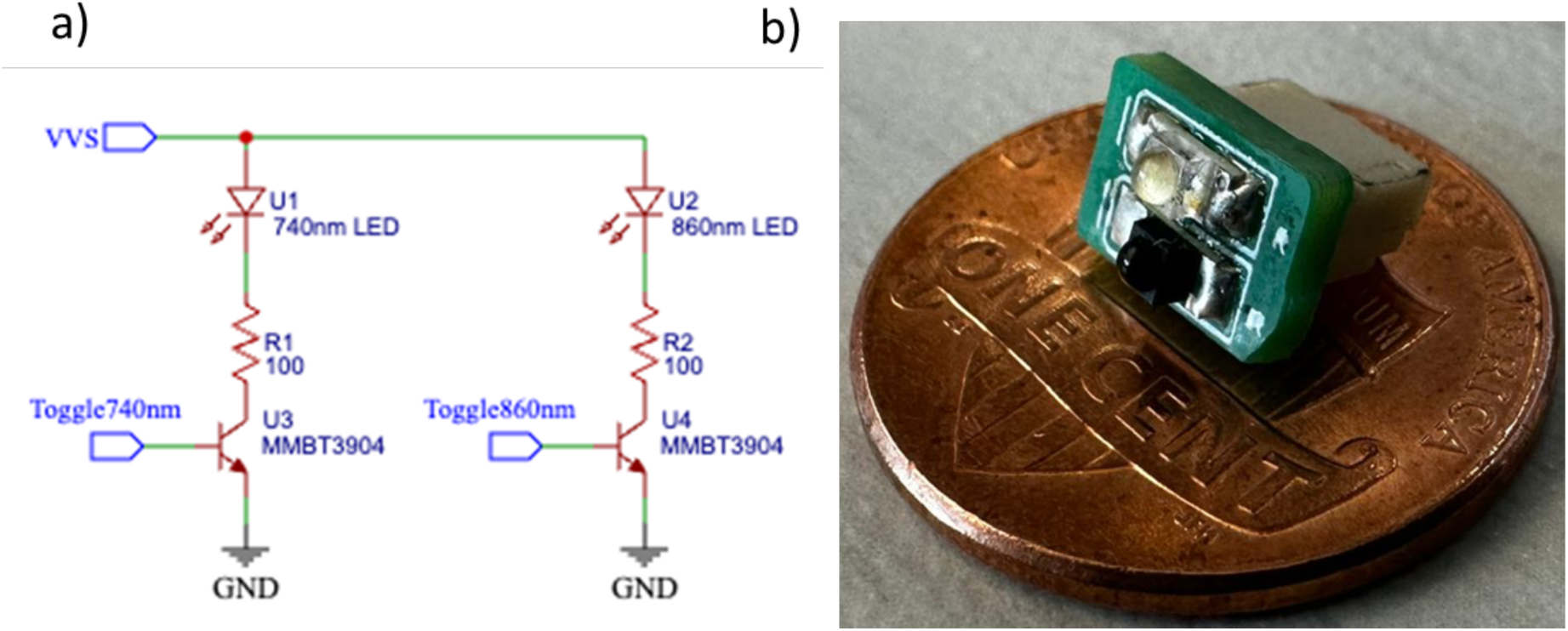
a) Schematic diagram of the dual-wavelength emitter circuit. b) Photograph of an assembled dual wavelength near-infrared LED emitter for fNIRS. The probe has been placed atop a USA penny for size reference.

#### 2.2.2. Detector design

The detector retrieves the near-infrared light backscattered from the tissue with minimal noise. It comprises a silicon photodiode (Vishay VBPW34S) with a large sensing area (7.5 mm^2^), wide viewing angle (±65°), rapid response time (100ns), sensitivity to the near-infrared wavelengths (600 nm to 900 nm), and linear response 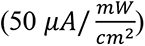. A Texas Instruments LMV793 rail-to-rail trans-impedance operation amplifier converts the current output of the photodiode into a voltage and is configured with a responsivity of 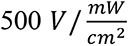. A Renesas Electronics ISL21080 precision voltage reference chip is also integrated to add a 0.9V DC bias voltage to the measurement. The detector requires supply voltages between 3.3 V to 5 V to operate correctly, and it includes decoupling capacitors for both the supply and voltage reference signals to minimize detector noise. Like the emitter, the detector has been packaged into a two-layer, double-sided PCB with a 4-pin JST connector. Schematics of the light detector are shown in Figure 3a, and photographs of the assembled detector are shown in Figure 3b. Unlike the emitter, all detector circuitries are mounted onto the PCB.

**Figure 3:**
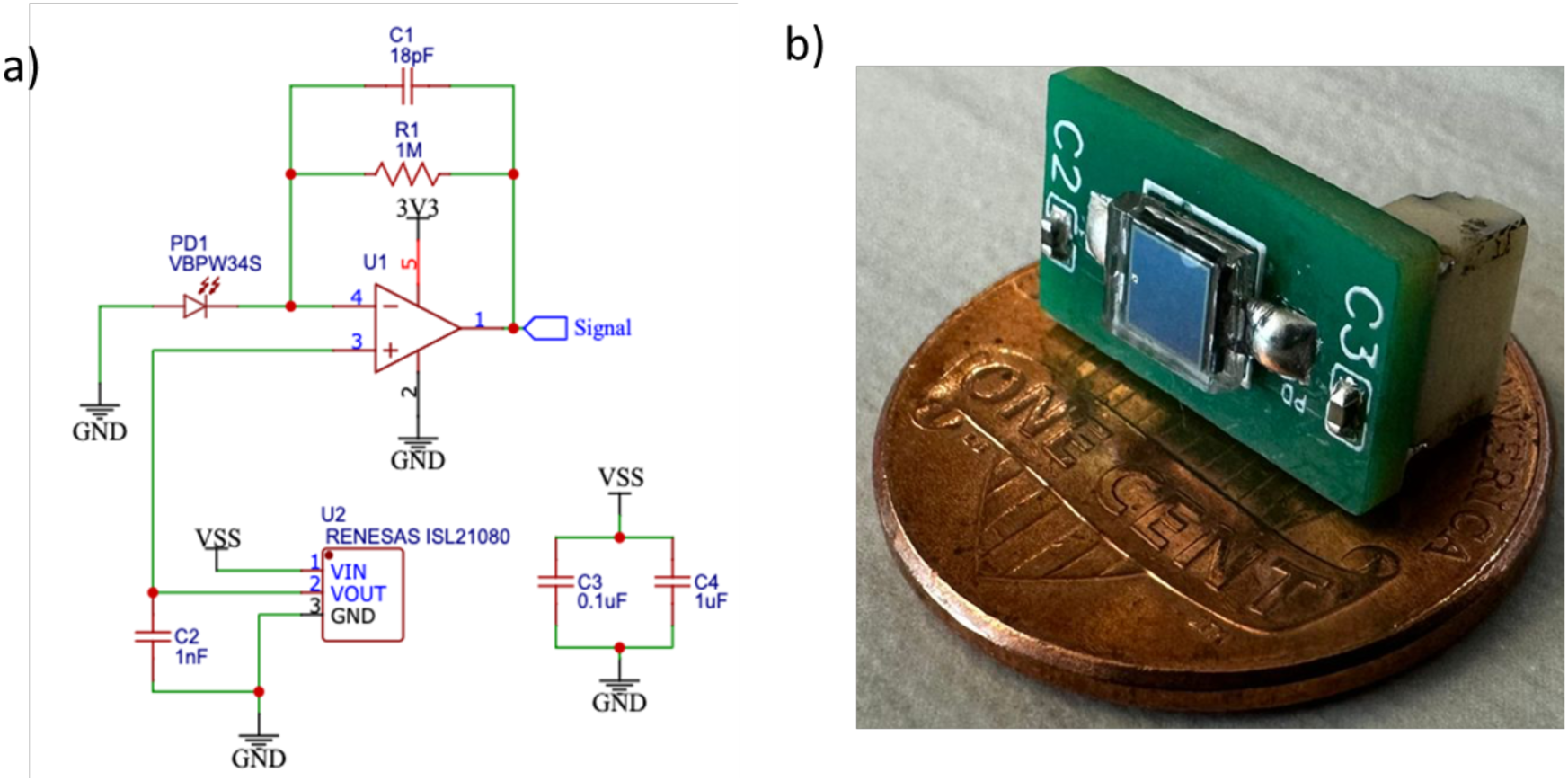
a) Schematic diagram for the light detector circuitry. b) Photograph of an assembled light detector. The probe has been placed atop a USA penny for size reference.

#### 2.2.3. Controller design

The controller board comprises four subsystems: 1) power management, 2) emitter control, 3) detector control, and 4) data aggregation and wireless communication. The block diagram of the controller board and subsystem connections is shown in Figure 4a.

**Figure 4:**
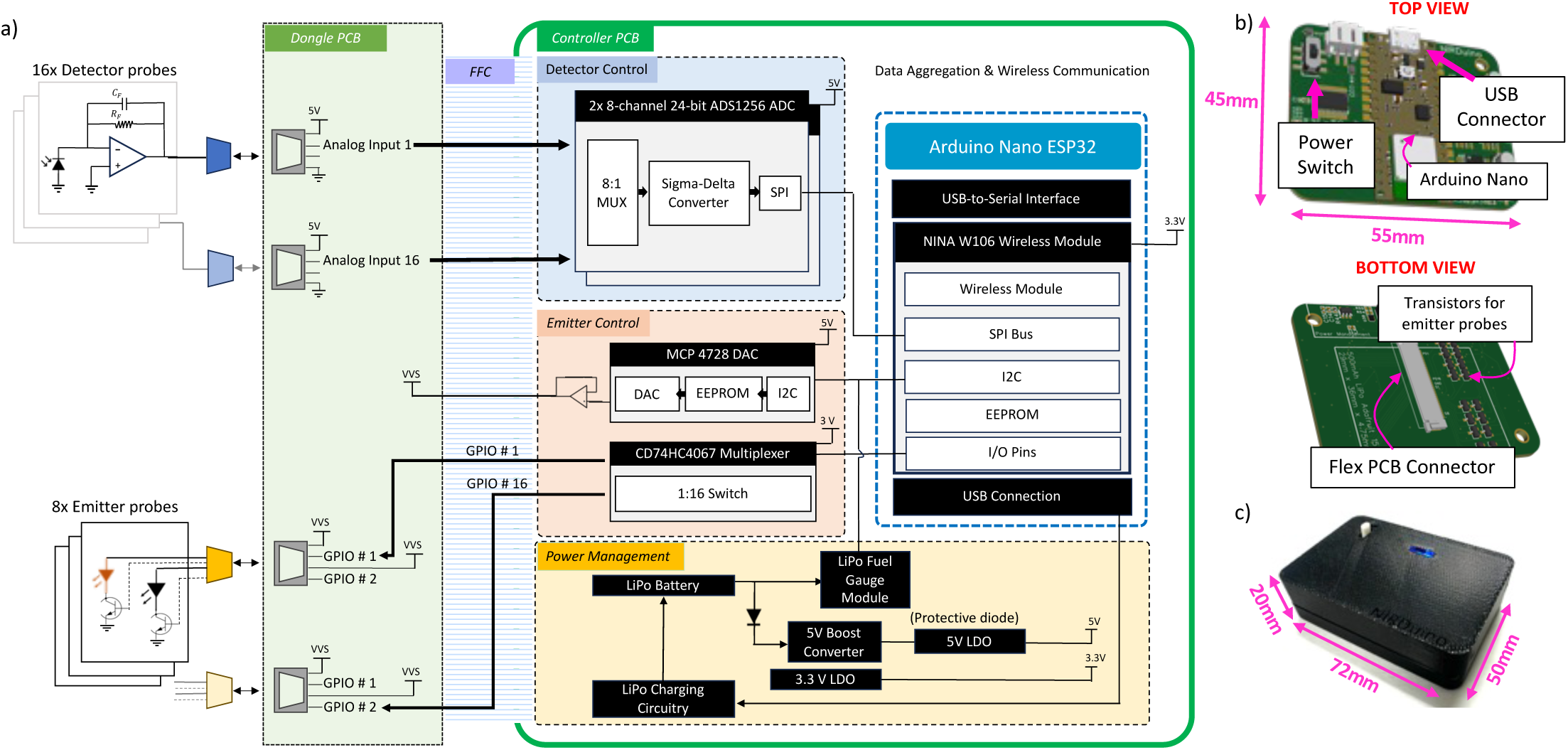
a) System block diagram of the NIRDuino electronics. Starting from the left, individual emitters and detectors are connected to a dongle with 8 emitter sockets and 16 detector sockets. The dongle is connected to a controller PCB with a flat flex cable. Detector control, emitter control, power management, data aggregation, and wireless communication are all housed in the controller PCB. b) Top and bottom views of the 3D render of the central controller PCB. c) Controller enclosed within 3D- printed box. Enclosure includes a light guide for the multicolor LED onboard the Arduino® Nano ESP32 board

The power management subsystem (PMSS) retrieves a variable voltage (2.5V to 4.2V) from a lithium polymer (LiPo) battery and provides two stable voltage supplies of 3.3V and 5V to the rest of the subsystems. The 3.3V supply is achieved using a Texas Instruments TPS73630 linear- dropout regulator (LDO) and draws current directly from the LiPo battery. The 5V supply is achieved using a Texas Instruments TPS61023 buck-boost converter followed by a Microchip MCP1711 linear-dropout regulator (LDO). For battery charging, the PMSS has a Microchip MCP73831 that takes in a voltage supply from a USB and provides a controlled current supply to recharge a LiPo battery. For battery level monitoring, the PMSS has a Maxim MAX17043 chip.

The emitter control subsystem illuminates and controls the light intensity of up to eight emitters, each consisting of two separate LEDs. During operation, only one LED is illuminated at a time. Light intensity control is achieved using a VVC that can supply a voltage ranging from 0V to 5V to the LED; during the setup process, the researcher can select what voltage to supply to each LED. The VVC uses an MCP4728 12-bit digital-to-analog conversion (DAC) chip with a voltage follower circuit to ensure the stability of the LED driving currents regardless of the current draw. To turn each LED in an emitter on or off, a 16-channel multiplexer (Texas Instruments CD74HC4067) routes a digital “high” signal to the base pin on the emitter transistor’s base pin.

The detector control circuitry collects data from up to 16 individual light detectors. The analog signals from the 16 different light detectors are routed into two Texas Instruments ADS1256 analog-to-digital converters (ADCs). Each ADC can digitize the signal using a 24-bit delta-sigma analog-digital-converter (ADC) with an extremely low non-linearity of just 0.001%, an input-referred noise of 27nV, and configurable sampling rates up to 30kSPS. Both ADCs are provided a 2.5V voltage reference signal from an external Analog Devices ADR03 chip to ensure precision measurements.

For fNIRS data aggregation and wireless data broadcasting, emitter and detector control circuitry are managed using an Arduino® Nano ESP32 board. The Arduino® board has an onboard Electrically Erasable Programmable Read-Only Memory (EEPROM) module for storing configuration parameters and a multicolored LED used to alert researchers about connection/disconnection or low battery. Additionally, the board has a multicolored LED that serves as an indicator light for the controller. The ESP32 controls the emitter control subsystem using its I2C protocol bus and general-purpose input/output (GPIO) pins. It communicates with the light detector control subsystem using the serial-peripheral-interface (SPI) bus.

The controller PCB measures 55 mm x 45 mm. It has an onboard flex PCB connector (Figure 4b) and is housed in a 3D-printed enclosure with dimensions 72 mm x 50 mm x 20 mm (Figure 4c). The PCB enclosure includes a slot to let the light from the Arduino® board’s multicolor indicator LED pass through. To ensure this light is visible, the enclosure has a laser-cut, 3-mm-thick acrylic piece that diffuses the light.

The controller PCB has been made resistant to electromagnetic interference using two separate methods: 1) a ground plane has been included in the controller PCB, and 2) various dual decoupling capacitors have been utilized at the voltage supply input pins for all analog and digital ICs. The ground plane helps shield the PCB from external electromagnet interferences. Dual decoupling capacitors help take care of both large transient changes in current draw from the PWSS and high frequency interferences caused due to environmental interferences.

#### 2.2.4. Dongle circuit design

The flex PCB connector of the controller circuit is designed to interface with a separate “dongle” PCB (Figure 5) using a flat flexible cable (FFC). The dongle has 8 four-pin JST connectors to connect with and control up to eight emitters and 16 four-pin JST connectors to connect with and receive data from up to 16 light detectors. These JST connectors enable researchers to connect the dongles to individual emitters and detectors using cables marketed by Adafruit Industries under the brand name “Qwiic”, which are readily available.

**Figure 5:**
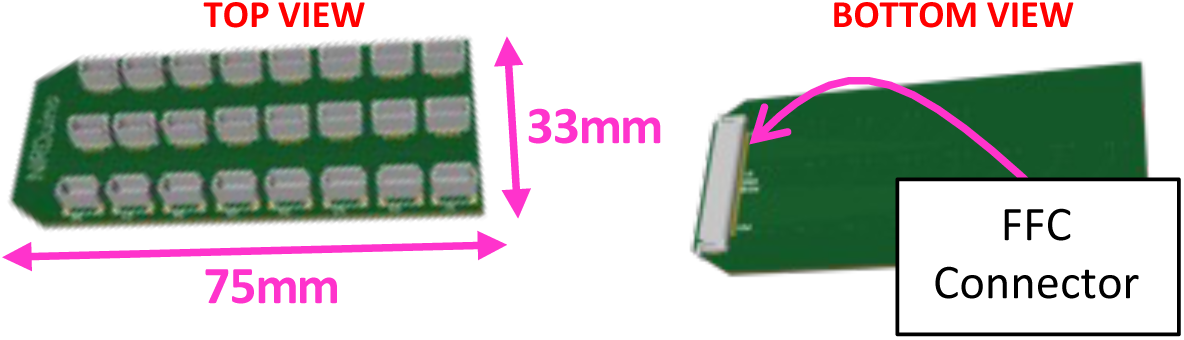
3D-render of the dongle PCB with a total of 24 JST-SH receptacles for each emitter and detector. The dongle connects to the controller board using a flex-PCB board.

#### 2.2.5. System assembly

Figure 6 shows a representative example of how the components interface together. The controller PCB (without enclosure) requires two connections: one to a rechargeable LiPo battery and one to the dongle circuit using a flat flex cable (FFC). The emitter and detectors interface individually with the dongle circuit using JST cables. The cable-based arrangement maximizes the ability of the researcher to adopt any emitter-detector layout they wish to utilize. (Note: The electronics assembly shown in Figure 6 is an actual assembly and not a 3D render, and the physical JST connectors are black in color, unlike their white appearance in the 3D render shown in Figure 5.

**Figure 6:**
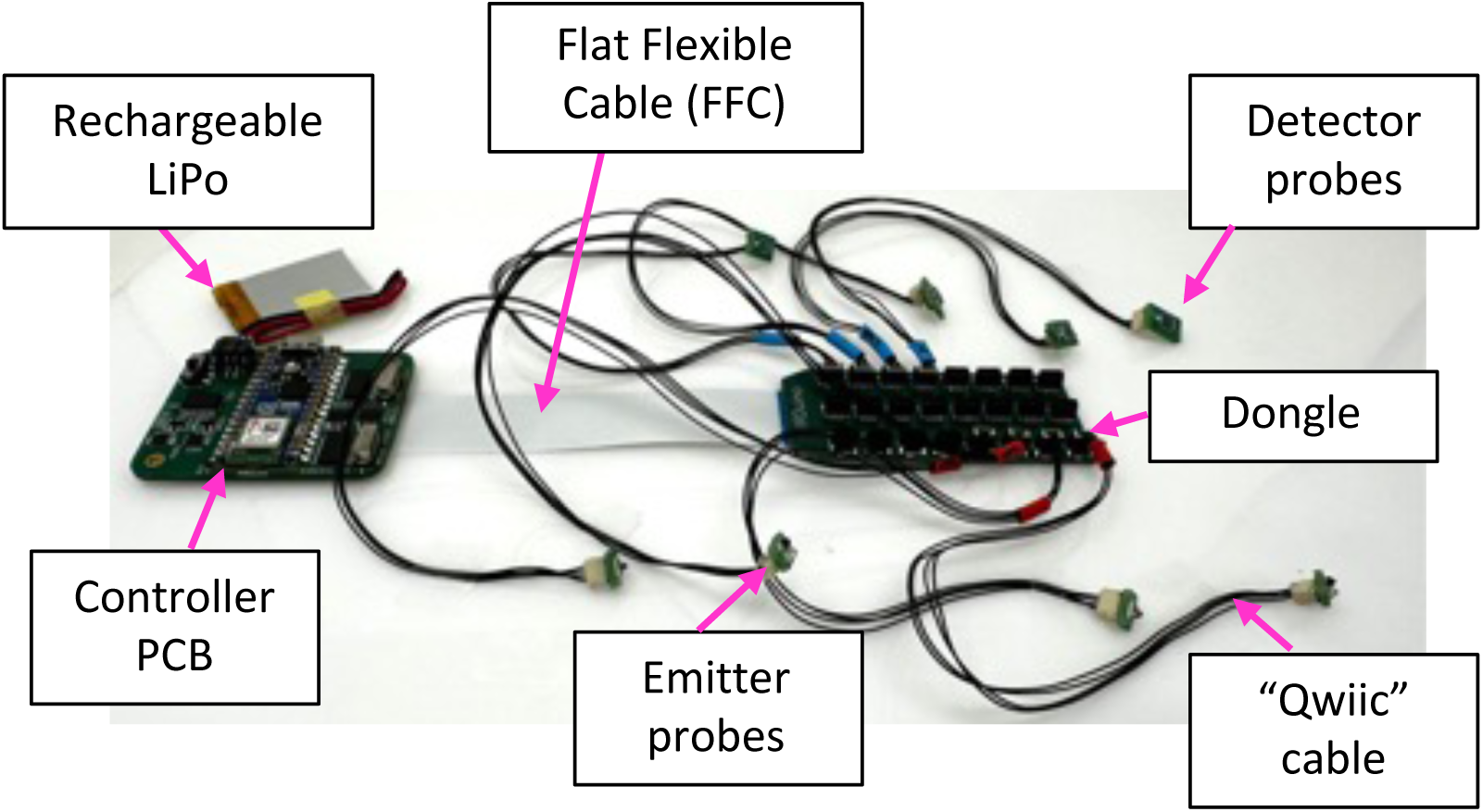
Photograph of the fully assembled electronics showing 4 emitters, and 4 detectors connected.

### 2.3. Software design

Two separate software are available for the researcher to easily set up their experiment, acquire and visualize the data, and store the measurements. Once installed on the devices, the software code does not need to be modified for unique experimental configurations and will enable fully mobile neuroimaging. The first software is designed to run on the fNIRS controller and is referred to as “firmware”. The firmware must be installed on the ESP32 microcontroller chip of the controller using a USB-A to USB-C cable. The firmware can simply be installed using a PC running the Arduino® integrated development environment (IDE) distributed by the Arduino® organization via their website. The second software is an Android^®^-based mobile tablet application. This application is responsible for wirelessly sending the experimental configuration to the controller, receiving data from the controller, and providing the researcher with data visualization and storage functionality. The controller and the mobile application were designed to wirelessly communicate with each other over Bluetooth Low Energy (BLE). In what follows, we first discuss the BLE communication, and then describe the individual software.

#### 2.3.1. Bluetooth communication and commands

In BLE configurations, “peripheral” devices (i.e., the controller) send data and “central” devices (i.e., the Android^®^ tablet) receive them. The peripheral stores and communicates data in a General Attribute (GATT) profile, which is a hierarchical data table organized around services, characteristics, and descriptors. Services are groups of data-storing variables called characteristics. Each characteristic stores a data value and can have several descriptors, which are additional meta- data describing the data values stored within the characteristic. There can be multiple services in a GATT profile, and each service can have multiple characteristics.

To achieve the desired data throughput and control requirements, the GATT profile for the NIRDuino system comprises a single service with three characteristics: one for communicating control configurations and two for communicating data. The control characteristic stores data for managing device configurations such as enabling/disabling data collection and the intensities of each LED of each emitter. This characteristic can be both read and written to by a “central” device, which in this case is an Android^®^ tablet. We found (through trial and error) that using two characteristics is the most effective way of sending the large volume of data from the controller to the Android^®^ tablet at the required data rate of approximately 5 Hz. Each data communication characteristic is set up to “notify” the Android^®^ tablet each time its value is updated. The values contain timing data, fNIRS data and battery level measurements, and they update whenever the controller collects data for any emitter-detector combination during fNIRS measurements.

The complete block diagram showing the flow of data between the NIRDuino system and the mobile application using the GATT profile is shown in Figure 7. Both the firmware and software can asynchronously read and write to specific characteristics of the profile. However, by design, BLE only permits one read/write command to be sent by either device at a time.

**Figure 7:**
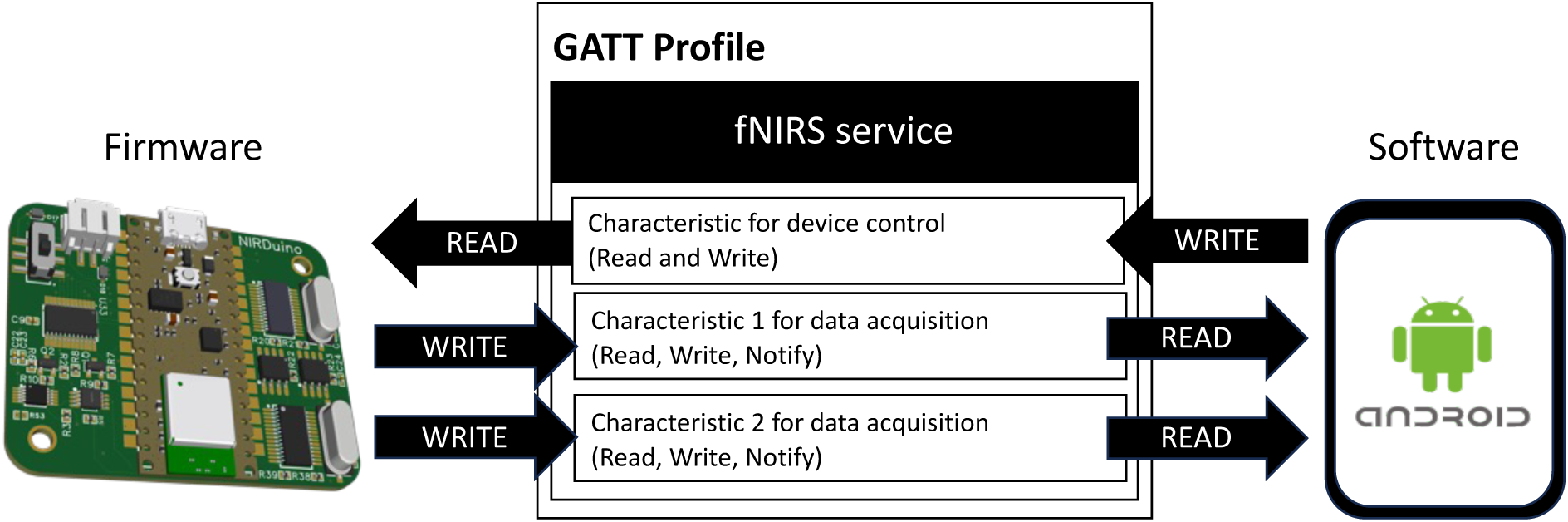
Data flow between the NIRDuino controller and the mobile device as defined by the Bluetooth Low Energy GATT profile.

#### 2.3.2. Firmware design

The objective of the firmware is to enable fNIRS data collection and broadcast the data over BLE. The timing diagram of the data collection process is shown in Figure 8a. Each round of fNIRS data collection comprises 33 cycles. The first 16 cycles illuminate the LEDs in each emitter one at a time; in each case, the 740nm LED is illuminated first, and then the 860nm LED. The LED power levels for these cycles are set with the assumption that these are “long” channels, with emitter-detector pair distances ranging from 20mm to 40mm. During the next 16 cycles, we repeat the process of sequential LED illumination, but this time at a lower power, as we assume that these are for collecting “short” channel data for emitter-detector distances below 20mm. Finally, we run a dark-measurement cycle where all LEDs are turned off and all 16 detectors values are measured sequentially, as before. Each cycle lasts roughly six milliseconds (ms); hence, it takes 198 ms to collect a full dataset. Raw voltages from all 16 light detectors and the elapsed duration of each cycle (in ms) are stored in a large data array.

**Figure 8:**
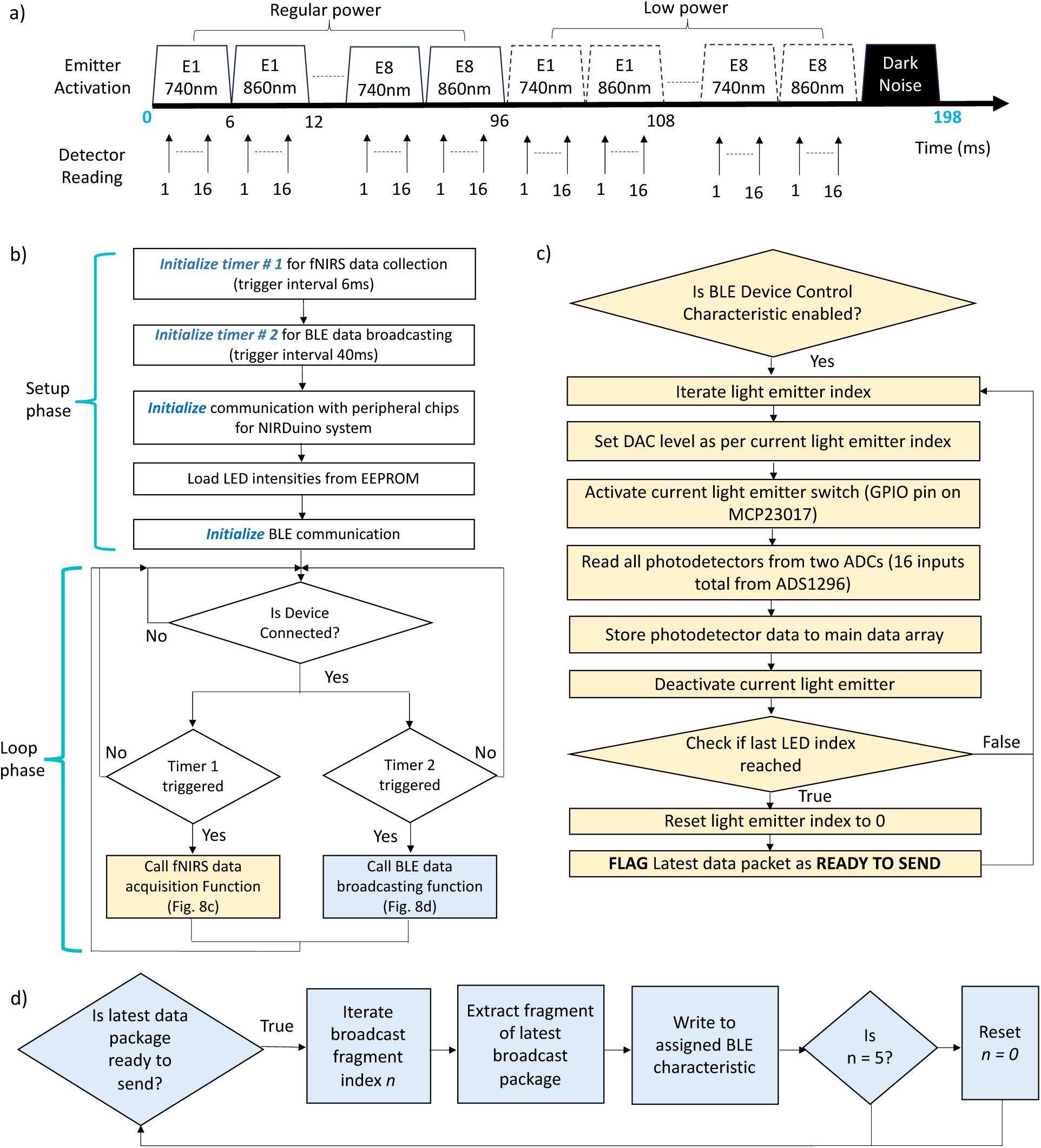
a) Fixed timing diagram for fNIRS measurements regardless of the chosen emitter-detector layout. b) Overall state machine diagram for Arduino®-based NIRDuino controller firmware. There is a setup phase and a loop phase wherein the loop phase is used to manage timer triggers. c) State machine of function triggered by timer assigned for fNIRS data collection. d) State machine of function triggered by timer assigned to BLE Data broadcasting.

The firmware is written using the easy-to-use Arduino® integrated development environment (IDE). Like any Arduino® firmware, there is a setup phase that prepares the electronics for all operations and then there is a loop phase that performs the same operations repeatedly. The overall state machine diagram for the firmware is shown in Figure 8b. The setup phase of the firmware involves setting up two timers. Timers are operations designed to trigger repeating operations at precise intervals of time. The two timers used in this firmware are for fNIRS data collection (Timer 1, interval = 6ms) and BLE data broadcasting (Timer 2, interval = 40ms). Next, the Arduino® board initializes communication with peripheral chips such as the ADS1256 chip used in the NIRDuino system. Following this, previously used LED intensities for fNIRS data collection are loaded from the EEPROM (i.e. the firmware remembers the last LED intensities used). The setup phase concludes with initialization of BLE communication. The name of the device is hardcoded to “BBOL NIRDuino (ESP32).” However, if the researcher wishes to change the name, it can be easily done by editing the relevant line in the firmware code in the Arduino® IDE. The only constraint is that to use NIRDuino with the provided mobile app, the new device name must have the keyword “BBOL” in its name. Once this setup process is done, the firmware enters the loop phase. In this phase, the firmware continuously checks if the device is connected to a BLE device and checks when the timers are triggered. To achieve fNIRS data collection, the firmware uses the state machine shown in Figure 8c each time Timer 1 is triggered. To simplify the control protocol and enable versatility, the firmware loops through every emitter-detector pair possible. Once all data have been collected and packaged for broadcasting, the firmware broadcasts the BLE data in five fragments of the output data package every 40ms (Figure 8d), or whenever Timer 2 is triggered. Hence, the cumulative BLE data rate is ∼5 Hz (or a period of 200ms). Regardless of how many emitters or detectors are selected for use in the neuroimaging study, all LEDs and detectors are cycled through; hence, the data rate is held constant. This approach ensures that the researcher can use the same NIRDuino firmware for any emitter-detector layout regardless of whether they use all or a subset of the available emitters of detectors.

One challenge with BLE communication is that it is an asynchronous communication technology. To accommodate this limitation, alongside the fNIRS data, the elapsed duration for collecting data from each emitter is also sent over BLE. This helps ensure that any variations owed to delays (and subsequent speed-ups) during BLE data collection over the wireless interface are circumvented and only timestamps associated with actual data are collected.

#### 2.3.3. Mobile app design

The Android^®^ mobile application was programmed using Android^®^ Studio and tested on a Samsung Galaxy Tab A7 Lite device (8.5” display) and a Google Pixel 3 smartphone device (5.5” display). To use this application, a packaged binary (“.apk file”) is available for download from the OSF repository, and the user will not need to use Android studio themselves. The .apk file can be loaded to any Android device for which the user has enabled developer status, which is a simple process one can learn to do with online resources.

The application comprises two screens: a device collection screen (Figure 9a) and a data acquisition screen (Figure 9b). Note: the controller must be turned on before using the app, and the researcher needs to ensure that the multicolor LED onboard the controller has turned blue. Conveniently, the controller will be ready to connect regardless of how many emitters or detectors are attached.

**Figure 9:**
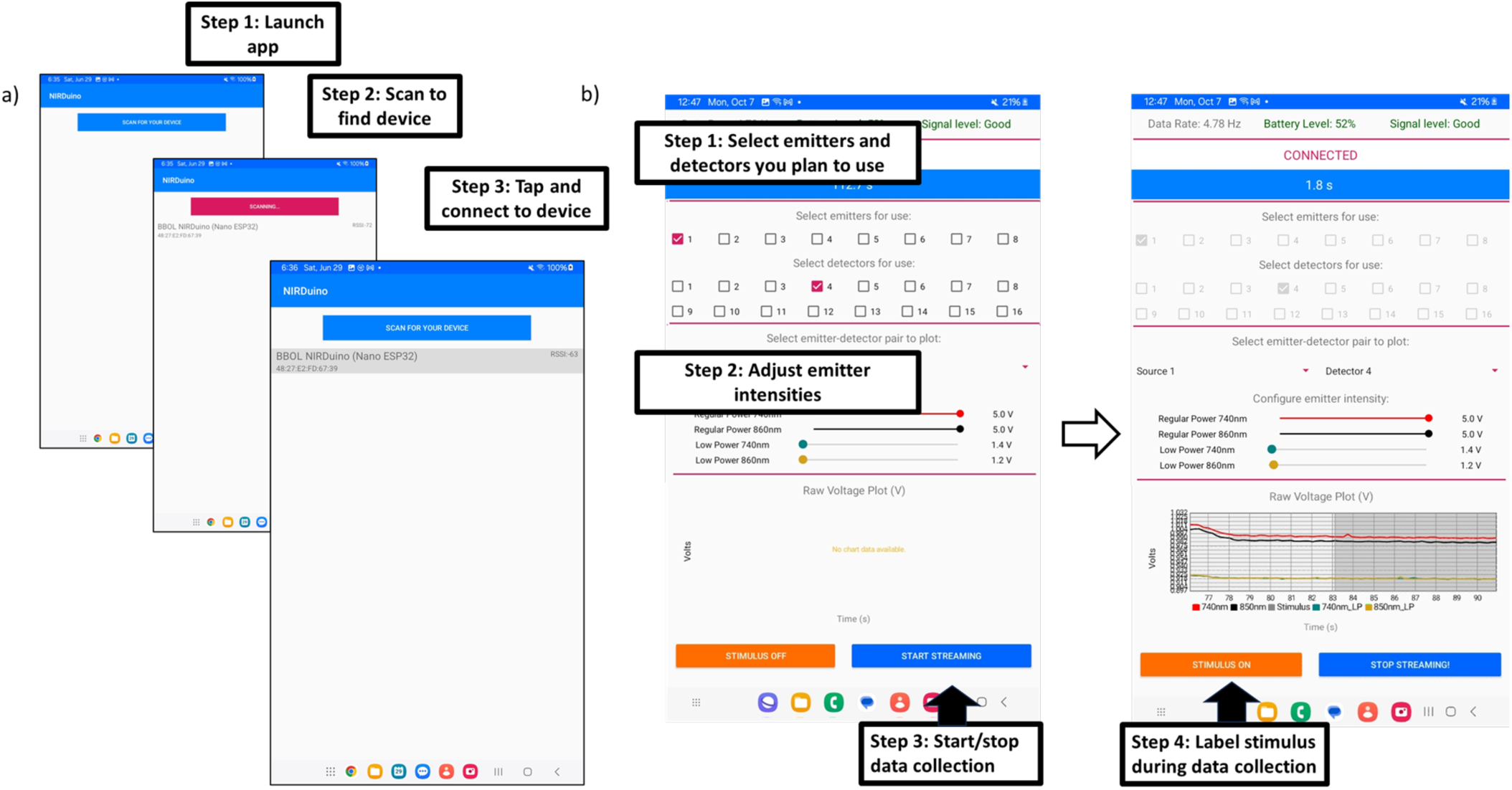
a) The device selection screen has three steps. The researcher should launch the app, tap a button to scan for nearby devices, and then select a device from a list of available options. b) The data acquisition and configuration screen of the mobile app. The first step of the app is selection of the emitters and detectors connected to the dongle for the experiment. Second, the LED intensities are adjusted for the experiment so that sufficient light reaches to detectors without saturation. To start data collection, a simple start/stop button is provided to the right. When data collection has started, stimulus periods of the data can be labeled on/off using the button on the left.

The device selection screen is the first screen of the app and helps the researcher find, select, and connect to their NIRDuino device (Figure 9a). When the app is launched, the screen consists of just a blue button on top of the screen. The researcher can tap this button to scan for and find nearby BLE devices (the button changes color during the scanning process to indicate when scanning is taking place). After scanning is complete, a list of available BLE devices (devices must be turned on) is provided to the researcher. To simplify the selection process, the list has been filtered to look specifically for the devices with the phrase “BBOL” in the name. To connect to a device on the list, the researcher just needs to tap the desired device on the list and the app will automatically proceed to the data acquisition step. If devices are not renamed in the provided firmware, researchers are recommended to only turn on one NIRDuino device at a time. The mobile app can differentiate between and separately list multiple NIRDuino devices based on their MAC addresses, but this is unintuitive for the researcher.

The data acquisition screen (Figure 9b) allows the researcher to configure their NIRDuino system for their experiments. The top of the screen presents information about the connection status, data rate, battery power level, and signal quality of the BLE connection (based on received signal strength indicator or RSSI value). An RSSI level greater than -55dBm (corresponding to 90% signal quality) was selected as the threshold for “good” signal quality on the appsig^61^ This RSSI level is typically considered sufficient for real-time data transfer^62^. The section below that enables them to simply select which emitters and detectors they wish to use (Figure 9b, step 1) for their experiments and to adjust the intensities of the emitters (Figure 9b, step 2). Intensity adjustment may be necessary to avoid saturation depending on varying tissue properties within and across individuals such as skin tone, subcutaneous fat, or muscle. The numbering on the emitters corresponds to the numbering of the connectors on the dongle. The “start streaming” button is located at the bottom right of the screen and initiates data collection once pressed (Figure 9b, step 3). During data collection, the app presents a multi-line plot of the data collected for the selected emitter and detector pair from the drop-down menu (Figure 9b, step 4). Each multi-line plot has four separate lines referring to the data from the two wavelengths in their regular and low-power illumination modes. During data acquisition, the researcher can label time windows as “stimulus” and “non-stimulus” using the “stimulus on/off” button. When the button is set to the “stimulus on” mode, a gray highlight shows up in the multi-line plot. Note that the stimulus labels are applied to all detector-emitter datasets, not just the one being viewed on-screen.

The fNIRS data for the selected emitters and detectors are continuously saved in a .csv file continuously after the “start streaming” button is pressed. Data collection stops when the button is pressed again. Data associated with unselected emitters and detectors are not saved. The stored data includes the timestamp of each data point collected, the stimulus status (on/off), the raw fNIRS data (comprising time stamps, regular power data for each emitter wavelength, and low power data for each emitter wavelength) and the LED intensities used to collect that data. The data are stored in the “Documents/NIRDuino” folder so that they can easily be accessed by the researcher by connecting their Android^®^ device to a PC with a USB interface. If the folder doesn’t exist, the app creates the folder prior to saving data to it. The filenames are saved with the format “YYYY_MM_DD_HH_MM_dataLog.csv”, where the time stamp refers to start of the data collection process.

### 2.4. System evaluation

#### 2.4.1. Phantom evaluation

The stability of the fNIRS signal was evaluated using a static tissue-mimicking phantom and a single emitter and detector pair. We used a static, tissue-mimicking fNIRS phantom with fixed absorption and scattering properties to ensure the measurements collected would only present instrumentation noise and drift. The phantom was prepared following the process introduced by Kawaguchi et al. ^63^, which involved homogeneously mixing and degassing a combination of epoxy, toner powder, and titanium dioxide. The mixture was cast into a cuboidal mold and sanded on all sides to dimensions of 62 mm x 37 mm x 17 mm. A 3-mm-thick sample of the final mixture was assessed with spectrophotometry using an integrating sphere, and the reflectance and transmittance measurements (λ = 600*nm* *to* 900*nm*) were collected using an Agilent Cary Series UV-Vis NIR Spectrophotometer (2-mm beam diameter). These data were post-processed using the inverse-adding doubling (IAD) model developed by Prahl et al.^64^ to calculate the absorption and reduced scattering properties shown in Figure 10a, and the values are consistent with phantoms published in the literature^63,65–69^. For evaluation, a single emitter and detector pair were over-molded with black-pigmented silicone to provide a spacing of 30 mm (a typical emitter-detector spacing for adults). The silicone-emitter-detector assembly was then placed over the phantom block (Figure 10b). The emitter and detector were connected to the NIRDuino controller, and the complete assembly was placed within a light-tight box to remove any ambient light. Firstly, the emitter was disconnected from the NIRDuino controller to turn off the emitter illumination and collect dark noise at the detector. The root mean square of the detector signal was collected and divided by the detector responsivity to calculate the minimum detectable light (noise equivalent power). The emitter was reconnected to the NIRDuino system, restarted, and the emitter was configured to its maximum power level of 100 mW/cm^2^. Data for the detector were collected for 30 minutes. The signal drift was calculated by measuring the % change in the RMS between the first and last 10 seconds of the data. The signal-to-noise ratio (SNR) of the signal was calculated by detrending the signal (i.e., subtracting the best line fit) and using the equation 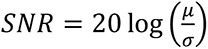, where μ is the mean of the signal and σ is the standard deviation over the last 10 seconds.

**Figure 10:**
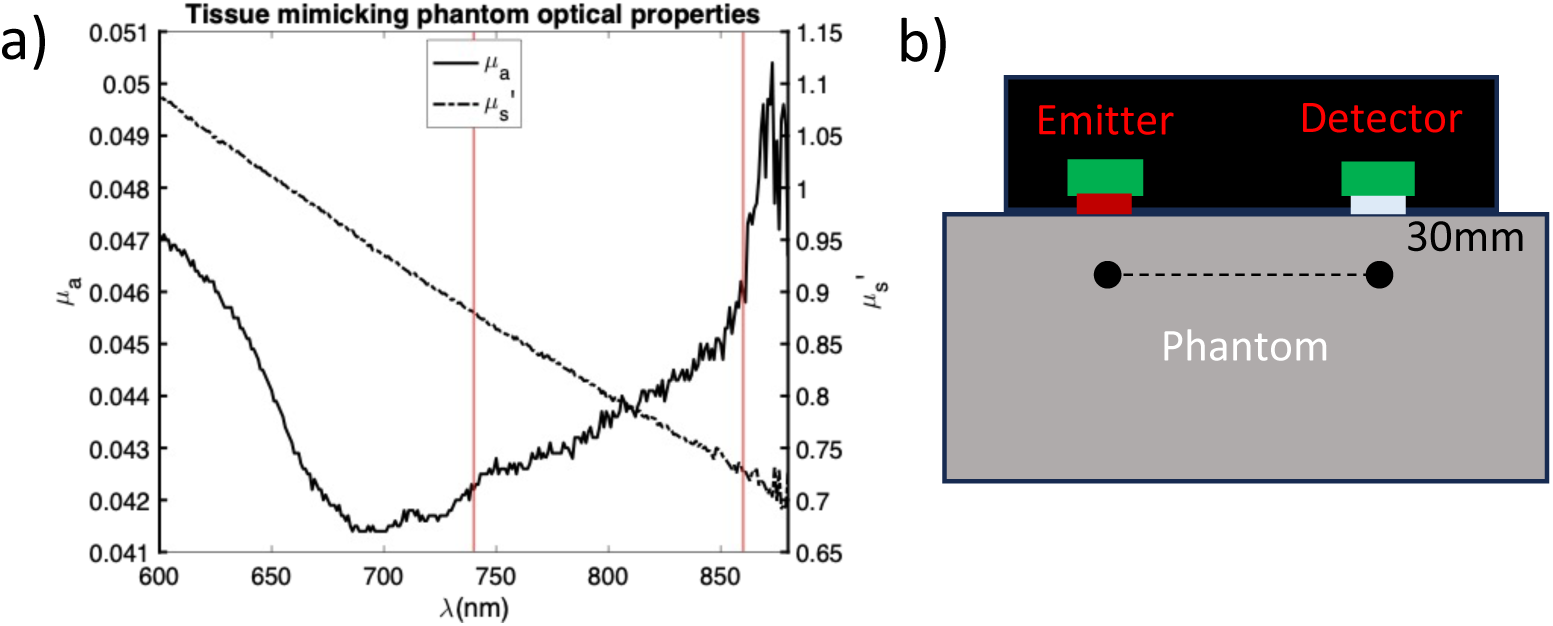
a) Spectrophotometer data for tissue-mimicking phantom. b) Visual representation of emitter and detector placed over a static phantom.

#### 2.4.2. Physiological Evaluation

Two physiological validation experiments were performed to demonstrate the Android^®^- configurability and modularity of the system: an arterial occlusion experiment on the forearm and an arithmetic experiment on the forehead^70–73^. The arterial occlusion experiment is a routine, non-cognitive experiment to evaluate the responsivity of fNIRS devices to an obvious change in oxy- and deoxy- hemoglobin. This experiment also provided an opportunity to demonstrate a non-brain measurement using the NIRDuino system. The arithmetic experiment is a cognitive experiment that was selected for its known activation in the left dorsolateral prefrontal cortex area accessible under the forehead. The forehead provides a hair-free window to the brain convenient for testing fNIRS systems such as NIRDuino.

Each experiment was conducted using a different emitter-detector layout; thus, a separate light-blocking interface was prepared for each experiment using molded silicone and laser-cut fabric assembled using a combination of stitching and adhesives. CAD files for both experiments have been shared. All data were collected wirelessly using the Android^®^ tablet. Notably, the setup process simply required rearrangement of the emitter and detector layout without need for any adjustments to firmware or software. A researcher on this team performed both physiological experiments independently to validate the functionality of the device. Given that the physiological experiments duplicate past experiments and the collected sensor data proves basic usability of the system, IRB approval was not required, and a single subject was sufficient for system evaluation. For the arterial occlusion experiment, three emitters and six detectors were arranged to measure three channels along the left forearm, placed downstream of a manually inflatable blood pressure cuff (mfg. by CVS). Each emitter was paired with one short detector placed 8-mm away and one long detector positioned at 30mm. The emitter-detector layout super-imposed over an assembled emitter-detector layout enclosed in light-blocking, blackened silicone is shown in Figure 11a. The complete enclosure consisted of a laser-cut pleather fabric glued to the silicone surface. To anchor the fabric-silicone enclosure to the forearm, additional straps with belt loops for tightening were sewn onto the fabric portion (Figure 11b). During the experiment, the blood pressure cuff was inflated up to 280 mmHg, held at constant pressure for 100 seconds, and immediately deflated afterward. fNIRS data were collected wirelessly during the cuff inflation process, during the cuff inflation hold, and during the release of cuff pressure.

**Figure 11:**
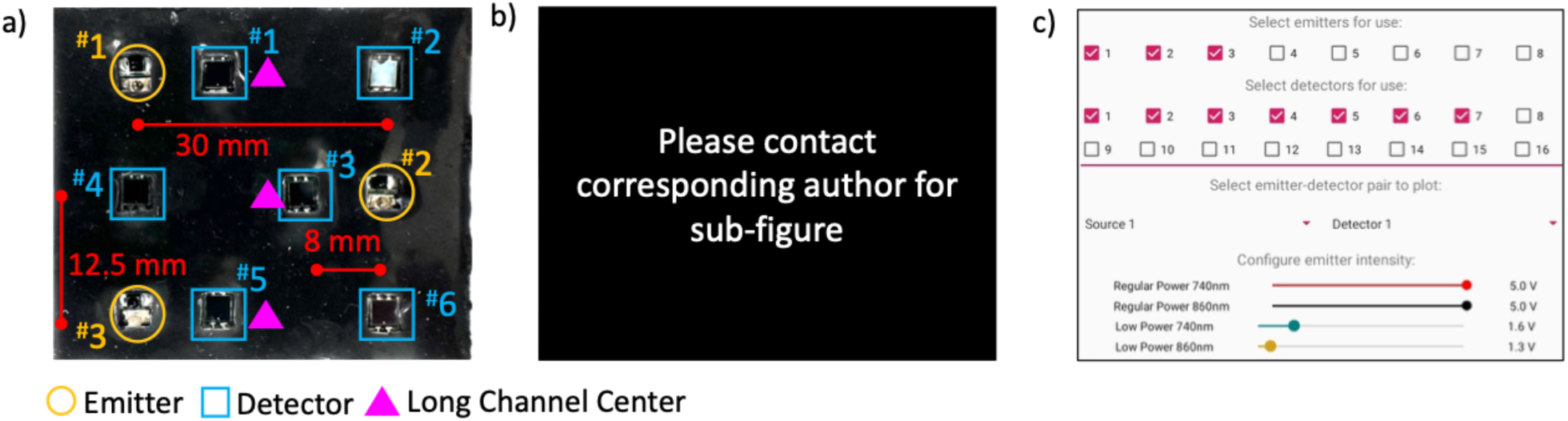
a) 3-channel emitter-detector layout for arterial cuff occlusion experiment. Emitter and detectors numbers are labeled using the “#” symbol. b) Placement of the layout on the left forearm. c) Segment of app screenshot for experimental setup.

For the arithmetic test, all 8 emitters and all 16 detectors of the system were arranged to measure 31 “long” fNIRS channels and six “short” fNIRS channels (a). The arrangement was designed to co-utilize multiple detectors for both “long” and “short” channel measurements. For “short” channel measurements, the detector signals are bound to saturate (i.e. reach the maximum measurable voltage of 5V) during regular power LED illumination. Hence, the second, low-power illumination mode ensures that the measurement can be conducted without saturating the detectors. An enclosure to attach the layout to the prefrontal cortex was created using plastic leather (“pleather”), blackened silicone, non- stretchy webbing, and plastic buckles. The bottom center of the emitter-detector layout (marked with an X) was aligned to the “Fpz” landmark on the forehead (***Figure 12***b). The mobile app was set to utilize all emitters and detectors as shown in ***Figure 12***c, and the LEDs were configured to maximum voltage (5V) for regular channel measurements and to lower values (*V*_740*nm*_ = 1.5V, *V*_860*nm*_ = 1.3V) for low-intensity cycles. The experimental protocol involved five cycles of a 30-second resting period followed by 30 seconds of arithmetic problem-solving. The arithmetic problem-solving phase involved the addition of two, two-digit numbers using mental calculations. The math problems were randomly generated by an app named “Mental Math Cards Games & Tips” published by Nicholas McNamara on the iOS store. The app was running on a separate device from the Android^®^ device used to collect the data.

**Figure 12:**
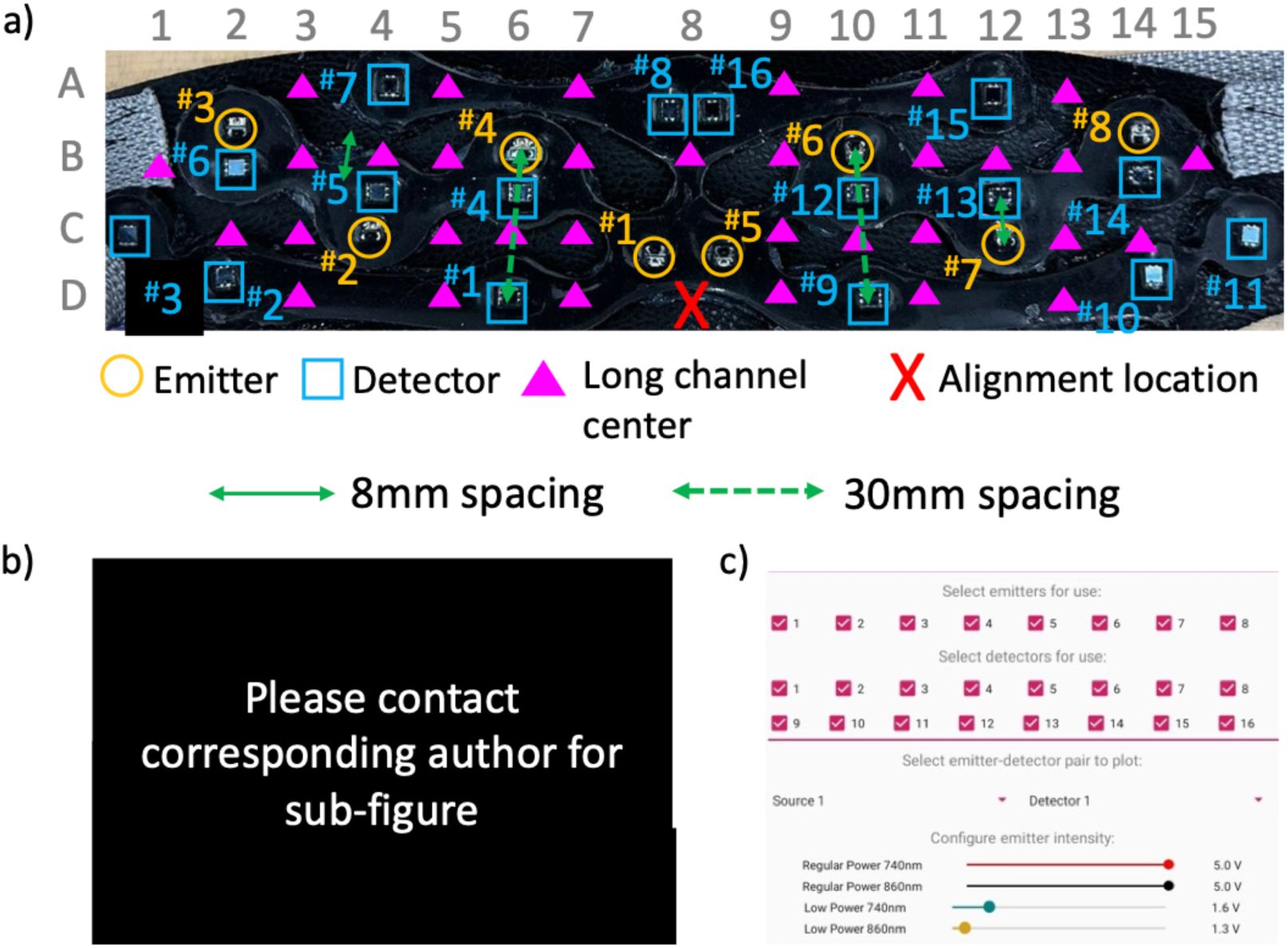
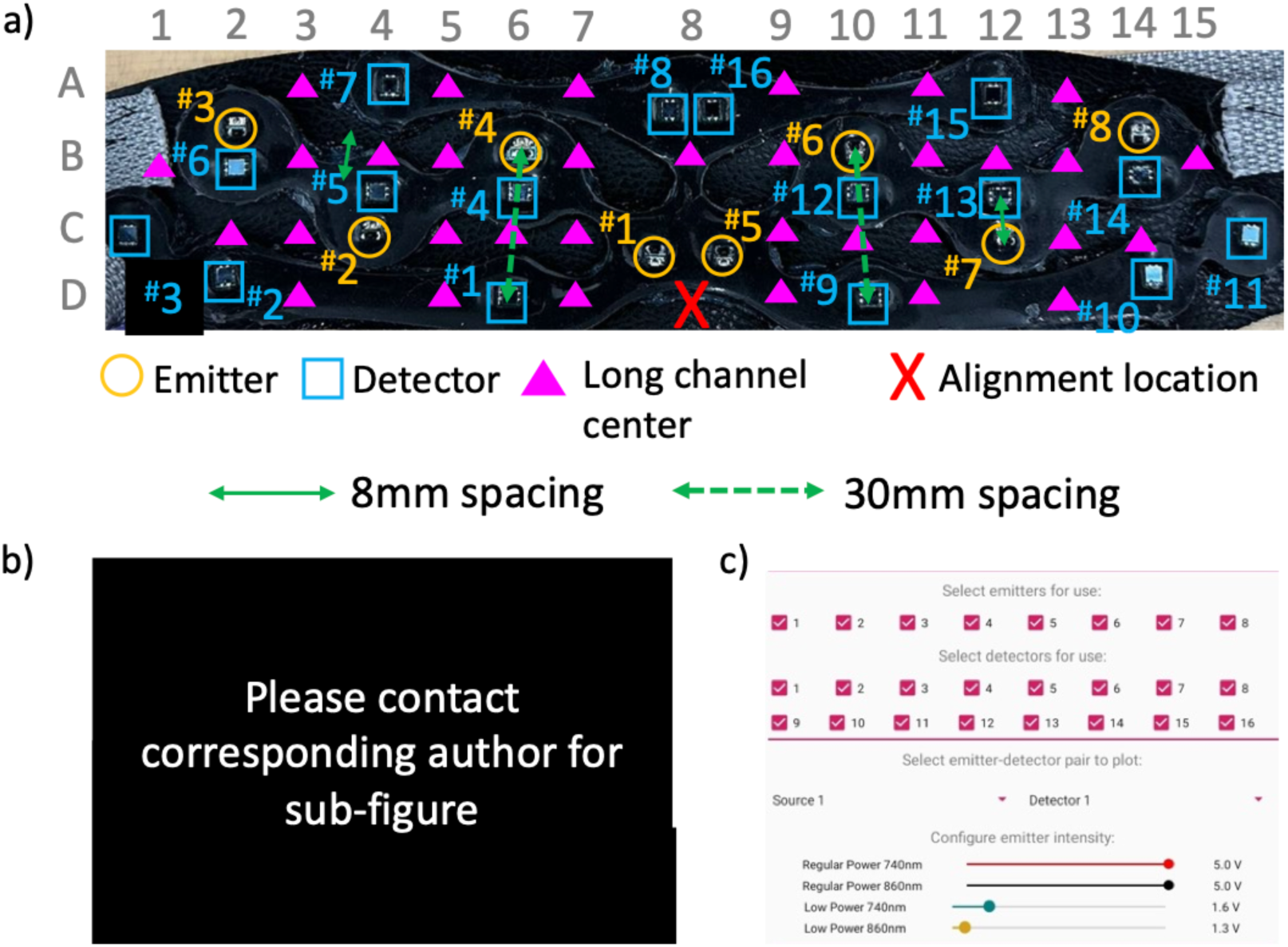
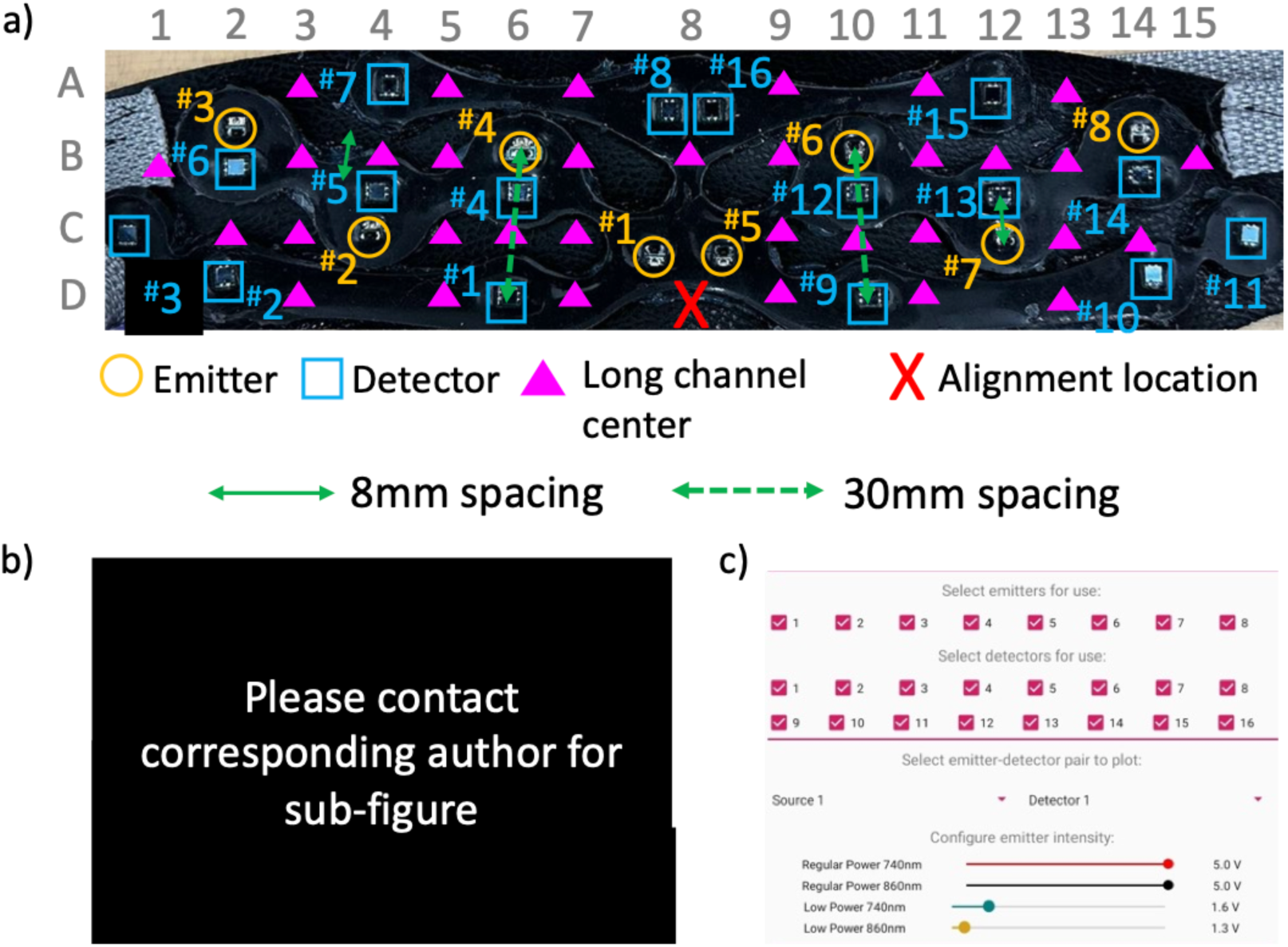
a) 31-channel emitter-detector layout for cognitive test measurements on the prefrontal cortex centered at site “FPz” at the “X”-marked alignment location. Emitter and detectors numbers are labeled using the “#” symbol. For easy referencing of the long channels formed by emitters and detectors spaced 30 mm apart (labeled with magenta triangles), row and column labels using letters and number (respectively) are provided using gray text along the vertical and horizontal axes. b) Placement of the device on the forehead centered at the “Fpz” landmark. The face of the individual has been completely concealed to avoid identification. c) Mobile app setup for data collection.

#### 2.4.3. Data processing

After experimentation, all the raw voltage data from the two experiments were retrieved from the mobile device to a desktop computer for subsequent analysis using MATLAB. To convert raw data into changes in hemoglobin measurements, all data were first median filtered (order = 21) to remove any noise and then converted into optical density values using equation (1) below, where V_λ_*_,_* is the voltage measurement for each wavelength and μ is the mean of the signal. The optical density process normalizes all intensity measurements according to the coupling and skin tissue properties (such as melanin content of the skin) of the emitter/detector with the scalp.

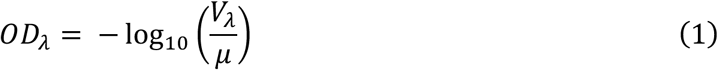

The optical density values were then converted to oxy- and deoxy- hemoglobin measurements using equation (2).

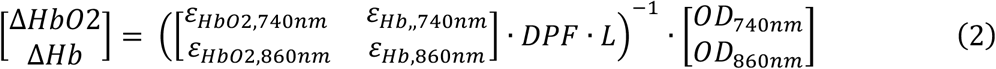

The extinction coefficients 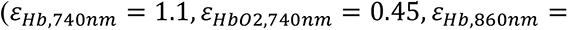 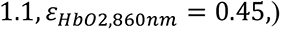 for both experiments were retrieved from work done by Zhao et al ^74^. Diffuse path-length factors (DPF) of 4.38 (*λ* = 744*nm*) and 3.94 (*λ* = 833 *nm*) for the forearm were utilized from work by Duncant et al.^75^ to estimate hemoglobin measurements. Although these values do not equal the LED wavelengths utilized for this study and may not accommodate for the adipose tissue content in the forearm used, they provide close approximations for our demonstration purposes. Precise, wavelength-specific DPF values for the forearm may be achieved using a combination of simulation and ex-vivo tissue spectroscopy, but that is out of the scope of this work. Wavelength- and age-specific DPF values of 6.25 (*λ* = 740 *nm*) and 4.89 (*λ* = 860 *nm*) for the forehead were calculated using an age- and wavelength- specific equation derived by Scholkmann et al^76,77^. These post-processed values were then detrended (subtracted the best straight-fit line), and bandpass filtered between 0.01 to 0.9 Hz ^16,78^ to minimize non-neurological physiological noise (cardiac, respiratory, blood pressure, and Meyer waves) or instrumentation noise (motion artifacts, electromagnetic interference). Finally, for each regular fNIRS channel (30 mm), data from the nearest short separation channel (8 mm) were normalized to the same amplitude ranges and subtracted ^79^. The final, post-processed signal was segmented into individual blocks, which were averaged to identify the block average for each emitter-detector pair. Each block included data from the resting period, stimulus period, and post-stimulus period. Since no classification was performed, no additional calibration was required.

## 3. Results and discussion

### 3.1. Phantom evaluation

The raw voltages from the photodiode during dark current measurements and voltage drift measurements are shown in Figure 13a and Figure 13b, respectively. The dark current measurement was utilized to calculate the noise-equivalent power by dividing the RMS of the signal by the configured responsivity (discussed earlier in “Methods”) of the signal. The noise equivalent power was found to be 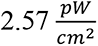 for the selected Vishay VBPW34S detector. A voltage drift of just 0.59% and 0.52% was found for the photodiode during 740nm and 860nm illumination, respectively. This is comparable to other published systems that have documented drift measurements of 0.3% (Wyser et al. ^80^), 0.56% (Liu et al. ^81^) and 0.42% (Lühmann et al. ^82^). The voltage drift experiment data were analyzed for their signal-to-noise ratio, which measured >50dB. SNR >40dB is sufficient for measuring hemodynamic response, and these results are comparable to past SNR measurements published 30mm emitter-detector spacing by Yaqub et al. (>50dB) ^83^, Liu et al. (<60dB) ^81^.

**Figure 13:**
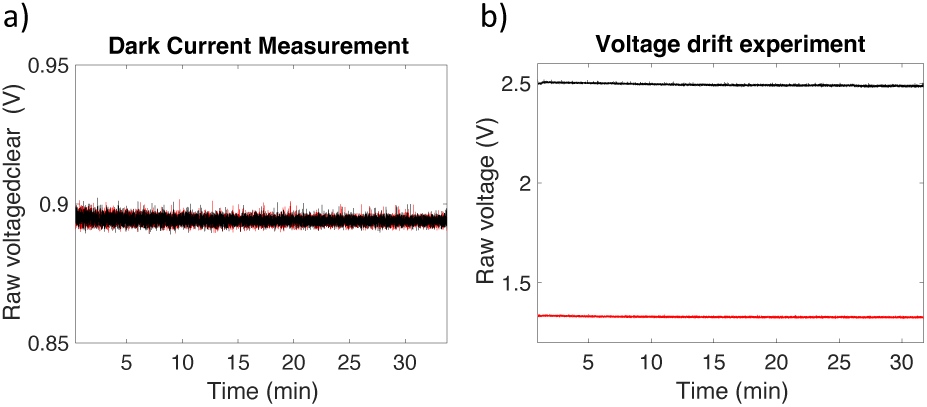
a) Raw voltage from photodiodes during dark current measurement for both 740 nm and 860 nm illumination periods. The LEDs were disconnected during this period. b) Raw voltage measurements from photodiodes for both 740nm and 860nm during illumination.

### 3.2. Physiological evaluation

The raw data from the arterial cuff occlusion experiments are shown in Figure 14a and Figure 14b, respectively, and the post-processed data are shown in Figure 14c. During the cuff occlusion period, the raw voltage for both wavelengths drops, demonstrating an increase in light absorption due to momentary local pooling of blood associated with the occlusion of any entry/exit of blood flow. In the post-processed data, this appears as a rise in oxyhemoglobin with a concurrent drop in deoxy hemoglobin. The cuff hold period of 100s resulted in a rise in the 860nm signal across all three channels alongside a drop in the 740nm signal. Finally, upon cuff release, there was a sharp drop in the raw 860nm signal accompanied with a rise in the 740nm signal. In the post-processed data, the cuff inflation period showed a sharp rise in oxy-hemoglobin and drop in deoxy- hemoglobin. These results successfully duplicate those from prior studies performing arterial cuff measurements ^83–86^.

**Figure 14:**
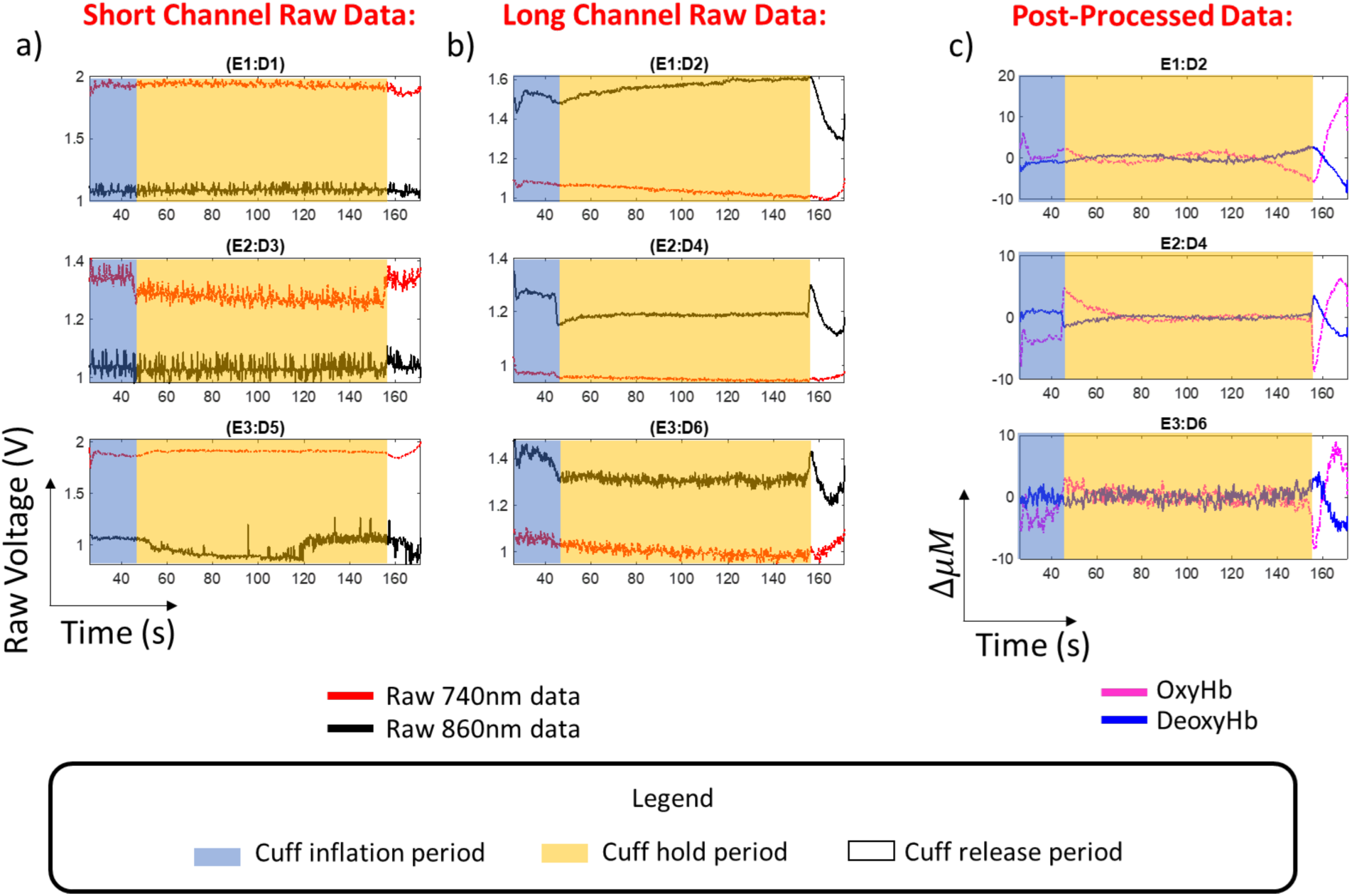
Results from arterial cuff experiment. a) Raw voltage measurements from short channels with emitters and detectors spaced at 8mm. Each emitter-detector pair shown in **Figure 11** has been labeled using the convention “E#:D#” b) Raw voltage measurements from arterial cuff experiment for emitters and detectors spaced at 30mm. c) post-processed data for arterial cuff measurement. These converted plots show the change in oxy- and deoxy- hemoglobin concentrations.

For the arithmetic test, the raw data for all “long channels” and “short channels” are shown in Figure 15a. The short channels are annotated using a bold magenta outline and were collected using the low power cycles during data collection to avoid detector saturation. The complete time series (271 seconds) of raw data for both 740nm and 860nm LED illumination of the forehead are shown in Figure 15a. A subset of raw voltage data from a long-separation channel associated with emitter 7 and detector 15 is shown in Figure 15b to demonstrate the clear presence of a plethysmography signal in the data, which indicates good signal quality. The post-processed, block-averaged data for the arithmetic test are shown in Figure 15c. Short channels are not shown in this plot since they have been subtracted from their nearest channels. Due to the larger number of emitter-detector pairs in this layout, an alternate channel naming method labeling rows with letters and columns with numbers has been adopted. Channels corresponding to the left dorsolateral prefrontal cortex region (B12, A13 marked using black arrows) were found to have a pronounced rise of ∼1μ*M* in oxy-hemoglobin and inverse drop in deoxy-hemoglobin compared to other regions of the prefrontal cortex. A zoomed-in view of channel B12 is shown in Figure 15d showing a sharp rise in oxy-hemoglobin from the resting phase during arithmetic testing (magenta arrow). The anti-correlated drop in deoxy-hemoglobin is marked using a blue arrow. A channel where no activation was observed has been shown in Figure 15e. This region-specific activation duplicates results published previously during past arithmetic experiments ^40,71–73^.

**Figure 15:**
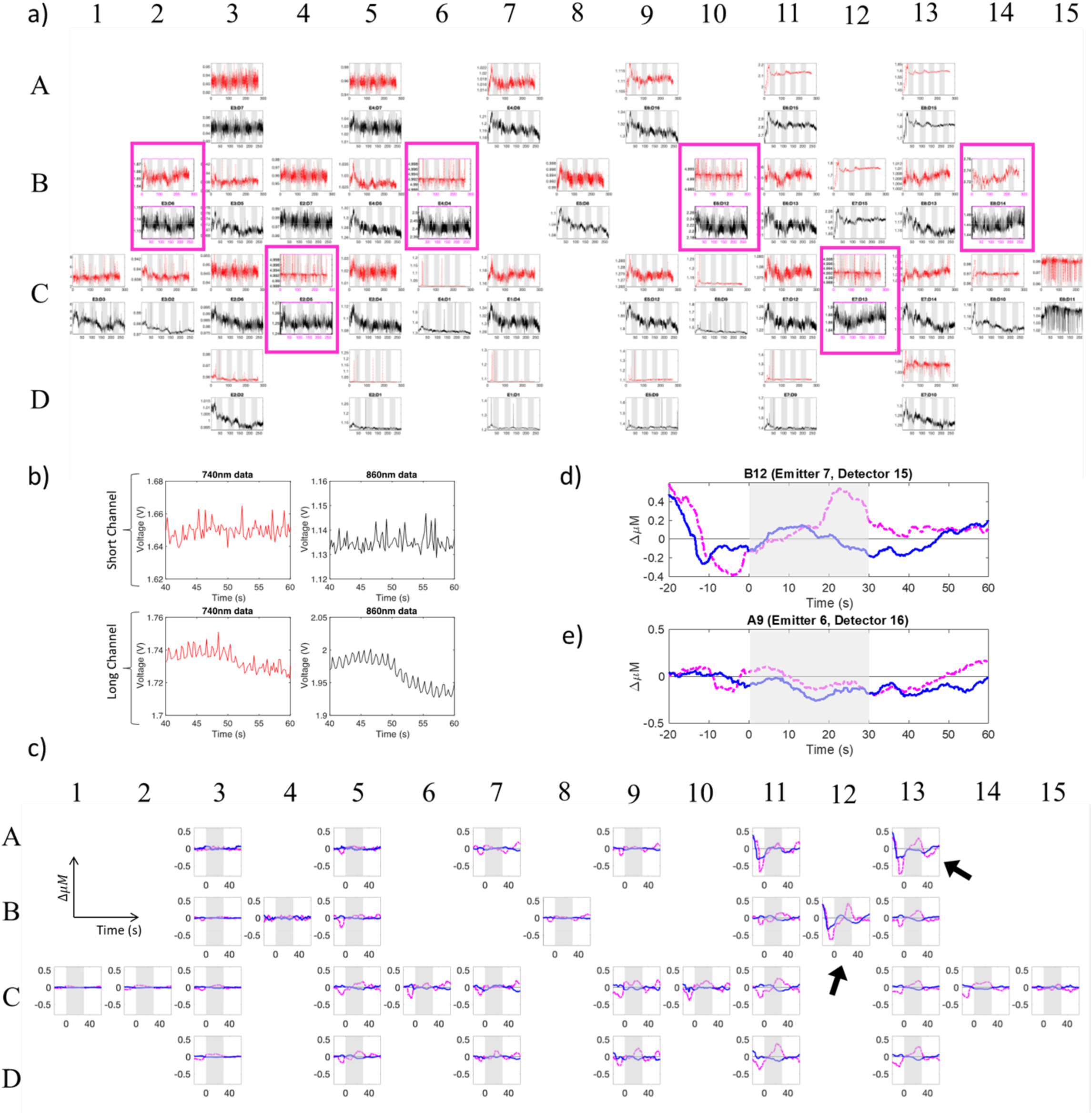
a) All raw voltage data for the arithmetic test. b) The plethysmography component visible in the fNIRS signal from the arithmetic test. c) Block-averaged post-processed data for arithmetic testing d) A zoomed-in view of channel B12 that shows a rise in oxy-hemoglobin during arithmetic testing like past studies. e) A zoomed-in view of channel A9 that did not show any response to arithmetic testing.

Results from both tests demonstrate that the NIRDuino system can be modularly configured for wirelessly monitoring changes in tissue hemoglobin concentrations using an Android^®^ tablet. No firmware or software changes were required to conduct these experiments, showcasing the versatility of the system for different types of measurements.

## 4. Conclusion

We have designed, tested, and made openly available the first Android^®^-configurable and modular fNIRS neuroimaging system. The system is very easy for researchers to adopt without any required engineering training, and costs less than $1000. As the methods and results demonstrate, the workflow of conducting any experiment with up to eight emitters and 16 detectors is as simple as checking a few boxes, and dual-intensity mode ensures that both long and short channels can be configured in layouts without worry of saturation.

There are still various opportunities for improvement. Firstly, the data rate of the system has currently been fixed to 5 Hz regardless of how many emitters and detectors are utilized. This can be overcome by updating the firmware to alter the fNIRS data collection interval based on the emitters and detectors selected for an experiment. Secondly the app platform can currently only be used to collect raw voltage data. For both conducting experiments and post-processing the data, additional software on other devices were required. These limitations can be overcome in the forthcoming years with adjustments to the NIRDuino firmware and app. Finally, the system requires use of a PC in order to assemble the hardware and program the software components. However, this shortcoming can be overcome through collaboration among researchers or by writing new software to program firmware onto devices using the mobile devices themselves.

The NIRDuino system offers the neuroimaging community significant improvements in affordability and usability. We hope that the community will use this device to build and share new datasets that lead to a more comprehensive, inclusive, and clinically actionable understanding of human cognition.

## Data Availability

All design, software, data, and post-processing files associated with this publication are available through an Open Science Framework repository found at this link: https://osf.io/ecy65/

## Acknowledgements

This work was funded by the Dorothy J. Wingfield Phillips Chancellor Faculty Fellowship and National Institutes of Health (NIH) grant # 1 R21 MH123873-01 and # 1 R21 AG072188-01. Teaching assistantship support for Anupam Kumar was provided by the Wond’ry Innovation Center at Vanderbilt University. We would also like to convey special thanks to Jeremiah Crosswhite (Vanderbilt Dept. of Arts and Sciences), Trieu Vy Truong (Vanderbilt Dept. of Computer Science), Lark Harrington (Dept. of Arts and Sciences), Allyria McBride (Vanderbilt Dept. of Arts and Sciences) for their technical support in sewing fabric enclosures for the fNIRS devices used in the papers.

